# The impact of Attachment Styles and Defense Mechanisms on psychological distress in a non-clinical young adult sample: a path analysis

**DOI:** 10.1101/2020.04.16.20055608

**Authors:** Giacomo Ciocca, Rodolfo Rossi, Alberto Collazzoni, Fiorela Gorea, Blerina Vallaj, Paolo Stratta, Lucia Longo, Erika Limoncin, Daniele Mollaioli, Dino Gibertoni, Emiliano Santarnecchi, Francesca Pacitti, Cinzia Niolu, Alberto Siracusano, Emmanuele A. Jannini, Giorgio Di Lorenzo

**Affiliations:** Department of Systems Medicine, University of Rome Tor Vergata, Italy; Department of Biotechnological and Applied Clinical Sciences, University of L’Aquila, Italy; Catholic University of “Our Lady of Good Council”, Tirana, Albania; Department of Mental Health, ASL 01 Avezzano-Sulmona-L’Aquila, Italy; Psychiatry and Clinical Psychology Unit, Fondazione Policlinico Tor Vergata, Rome, Italy; DIBINEM Department of Biomedical And Neuromotor Sciences, Unit Of Hygiene, Public Health And Biostatistics, Alma Mater Studiorum University Of Bologna; Department of Medicine, Surgery and Neuroscience, University of Siena, Italy; Berenson-Allen Center for Non-Invasive Brain Stimulation, Beth Israel Deaconess Medical Center, Harvard Medical School, Boston, MA, USA; Fondazione Santa Lucia, Istituto Di Ricovero e Cura a Carattere Scientifico, 00142, Rome, Italy, Braintrends Ltd, Applied Neuroscience, 00168, Rome, Italy

**Author notes:** Equal contribution (Giacomo Ciocca, Rodolfo Rossi).

**Keywords:** Psychopathology, psychological distress, Defense mechanism, Adult Attachment, Path Analysis

## Abstract

**Introduction:** Insecure attachment styles and immature or neurotic defense mechanisms are related to psychological distress. However, their mutual interaction in influencing psychological distress deserves further investigation.

**Methods:** One-thousand-one-hundred-twenty-nine University students were evaluated using the Global Severity Index of Symptoms Check List 90-Revised for psychological distress, Relationship Questionnaire for attachment styles and Defense Style Questionnaire for defense mechanisms. Following exploratory analyses, a Path Analysis was performed with psychological distress as outcome.

**Results:** Fearful and preoccupied attachment styles had a substantial impact on psychological distress. About 30% of their effect was mediated by Immature and Neurotic defenses, with the former having the major effect. Dismissing attachment showed no substantial effect on psychological distress. Secure Attachment and Mature Defenses had a small protective effect on psychological distress, but their inclusion in the path model did not improve overall goodness-of-fit. Attachment style and defense mechanisms accounted for nearly 25% of the variance in psychological distress.

**Conclusions:** The results showed that attachment styles and defense mechanisms have a substantial impact on psychological distress. The effect of attachment style is mediated by defense mechanisms. Individual differences in attachment style and defense mechanisms represent risk factors for psychological distress in young adults.

## 1 Introduction

Insecure attachment and immature or neurotic defense mechanisms are related to psychological distress. However, their mutual interaction in influencing psychological distress deserves further investigation.

The Attachment system is a regulator of infant-caregiver relationship and romantic relations in adulthood. Attachment theory postulates that emotions and relational behaviours in children and adults are driven by “Internal Working Models” (IWM) of the self and the others, arising from early relational experiences. IWM encode the beliefs about the self as being worthy of love and care in acceptable or unacceptable way, and about the others as trustworthy in providing care or being threatful (Bartholomew and Horowitz, 1991; Bowlby, 1973). In adults the *attachment style* (AS) is the pattern of relational expectations, thoughts, emotions, and behaviours resulting from IWM.

Adult attachment is organized along two orthogonal dimensions, avoidance and anxiety. Consequently, four styles of attachment are described. *Secure Attachment*, characterized by a positive model of self and others, with low levels of anxiety and avoidance; *Preoccupied Attachment*, characterized by a negative model of self and a positive model of others, associated with low avoidance and high anxiety. *Dismissing Attachment* with a positive model of self and a negative model of others, combines high avoidance and low anxiety. *Fearful Attachment* is based on a negative model of self and other, and it is associated with high avoidance and anxiety and high emotional dysregulation (Bartholomew and Horowitz, 1991; Mikulincer and Shaver, 2012).

Insecure attachment styles influence psychopathology (Fowler et al., 2013; Hazan and Shaver, 1987; Kobak and Bosmans, 2019), with preoccupied, fearful, and dismissing styles increasing the vulnerability to depressive and anxiety symptoms (Dagan et al., 2018; Manning et al., 2017; Marganska et al., 2013), personality disorders (Ehrenthal et al., 2018), psychotic diseases (Berry et al., 2007; Bowlby, 1988; Sheinbaum et al., 2014), and suicidality (Miniati et al., 2017). Conversely, secure attachment is more often found in healthy individuals with good interpersonal functioning (Karreman and Vingerhoets, 2012). As initially proposed by Bowlby, the risk for psychopathology is conveyed by negative IWM of self or others, or by strategies for processing attachment-related thoughts and feelings that compromise realistic appraisals. In a transdiagnostic model of Attachment and Psychopathology proposed by Ein-dor et al. (Ein-Dor et al., 2016), attachment anxiety hyperactivates coping strategies including upregulation of negative affectivity, hyper-vigilance to threat-related stimuli and distorted perception of others’ responsiveness. On the other hand, attachment avoidance involves deactivating strategies, including de-emphasizing threats, denying needs of care, and compulsive self-reliance. Therefore, attachment styles characterized by high anxiety could be associated with a more severe psychopathology compared to avoidant styles.

In psychopathology, adult attachment has a dual role. Firstly, as an intervening variable between early adverse experiences and psychopathological distress. Secondly, Attachment is a part of interpersonal cycles that mediates the effects of relational stressors on psychological distress, together with reasoning biases and defensive style (Kobak and Bosmans, 2019). In this regard, it is reasonable to expect that insecure attachment style could play an important role in psychological suffering together with some coping factors, including defense mechanisms.

Defense mechanisms are involuntary patterns of feelings, thoughts or behaviours that mediate response to stressful or threatening mental representations and feelings that would otherwise produce psychological distress, protecting the individual from mental suffering and one’s altered perception of self, others, or one’s own emotions (Vaillant, 2000, 2011). According to Vaillant (Vaillant, 2011), defenses share six important characteristics: 1) They ease the psychological distress arising from emotions and mental representations associated with inner conflict; 2) they are mostly unconscious; 3) they are distinct from one another; 4) they are dynamic and reversible; 5) they are not necessarily pathological, rather they can be adaptive, even creative; and 6) although they are invisible to the individual, they can be clear to the observer, appearing odd, even annoying. Research in the field of defense mechanisms have developed several classification systems, using both self-report scales and clinical evaluation methods (Draguns, 2004).

Defense mechanisms are organized along a gradient of personality development, from immature to mature defenses, with more immature mechanisms being associated with more severe psychopathology (Andrews et al., 1993). At the immature end of the spectrum, Psychotic and Immature defenses act through a massive reality distortion (such as delusional projection, psychotic denial, and psychotic distortion) or detachment from reality (autistic fantasy, dissociation). These mechanisms are associated with severe mental disorders, including psychosis and personality disorders, and lower interpersonal functioning, characterizing mood and severe anxiety disorders (Berney et al., 2014; Calati et al., 2010; Ciocca et al., 2017; Perry and Bond, 2012; Trower P, 1995), although immature defenses are normally present during childhood and early adolescence.

At an intermediate level, neurotic defense act essentially by keeping distressing thought content out of awareness, without affecting or distorting realty testing. Neurotic defenses such as undoing, repression, idealization and reaction formation, and are most common in personality disorders, anxiety disorders and depression and are normally present during adolescence (Vaillant, 2011). Finally, mature defenses are adaptive coping mechanisms that maximize gratification and allow relatively more conscious awareness of feelings, ideas, and their behavioural-related consequences. Mature defenses include sublimation, humour, anticipation and suppression, and characterize healthy adults, with higher self-esteem, extroversion, resilience and internal locus of control (Ciocca et al., 2015; Mickelson et al., 1997; Simeon et al., 2007; Treger et al., 2015; Vaillant and Vaillant, 1990).

Under a developmental perspective, immature and neurotic defenses are normally present during childhood and early adolescence (Vaillant, 2011), being gradually replaced by mature defenses during personality maturation. Among the potential factors that may influence the maturation of defense mechanisms, early relational experiences and attachment could play a key role.

In relation to attachment, defenses are conceptualized as mechanism that modulate the attachment system in order to reduce distressing feelings associated with negative expectancies, both at an *intrapersonal* and *interpersonal* level (Kobak and Bosmans, 2019), and are related to emotion dysregulation (Malik et al., 2015). For instance, dysfunctional defensive styles participate in the interpersonal cycles that convey the effect of attachment insecurity on psychological distress. Conversely, positive attachment experiences culminate in a secure attachment style that doesn’t need any reality-distorting defense (Cramer and Kelly, 2010). On the other hand, when attempts to gain support and care are unfulfilled, the attachment system can be deactivated, as in the case of avoidant attachment by more or less pathogenic defenses, for example denying attachment needs or denying one own’s weaknesses. Conversely, in preoccupied attachment, the attachment system is hyperactivated with desperate attempts to gain others’ proximity.

A connection between attachment styles, defense mechanisms and psychological distress in adults has been addressed in the literature by a relatively small number of studies. In one study, immature defenses and anxious attachment were associated with persistence of post-natal depression, although the interaction between the two factors was not tested (McMahon et al., 2005). Another research found a high prevalence of insecure attachment style and particular immature defenses among parents abusing their children (Cramer and Kelly, 2010).

Besharat et al. (2013) revealed a mediating role of defense mechanisms between insecure attachment and alexithymic traits in adolescents (Besharat, 2013), with insecure attachment being correlated with higher levels of alexithymia and secure attachment showing an inverse correlation. Laczkovics et al. investigated, in a considerably large sample of adolescents, a path model in which immature defenses mediated the effect of insecure attachment on psychopathology, suggesting that attachment style directly affects defense mechanisms, which in turn conveys the effects of insecure attachment on psychopathology (Laczkovics, 2018). In a study on healthy adults, the relationship between parental bonding, adult attachment and defense mechanisms was explored, with adult attachment style as dependent variable (Prunas, 2019). In this work, immature defenses were correlated with all dimensions of insecure attachment and inversely correlated to secure attachment, while secure attachment was found to be associated with mature defenses. Moreover, this study identified repression and splitting as defenses typical of avoidant attachment, while fantasy and projection were associated with anxious attachment. Our mediation model opens a relevant question about the direction of the effect between attachment and defenses. The authors specify a model in which defenses explain or predict adult attachment. This framework is reasonable considering that adult attachment as measured by the current psychometric instruments is a behavioural measure, regulated by implicit or sub-conscious factors such as defenses.

Under a theoretical perspective, it sounds reasonable that defense mechanisms could operate to “defend” a person from negative intrapsychic contents also derived from dysfunctional primary bonds. In this view in a putative pathway towards psychological distress, attachment styles would precede defense mechanisms. Hence, the aim of this study is to observe the relationship between attachment style and defense mechanisms and to assess their interaction with psychological distress in a large young adult sample using path analysis.

## 2 Materials and methods

### 2.1 Sample

For the current study, we performed a cross-sectional study of two samples from Italy and Albania, with a final sample size of 1129 subjects.

The first sample is a convenience sample of 551 university students recruited at the University of L’Aquila between 2014 and 2015, in the context of a larger cohort study. Five hundred and eighty students were recruited at the beginning of university classes of all university faculties, and they were asked to fill in a paper and pencil questionnaire on site after providing informed consent. No incentives were provided to the participants. Of these, 18 did not provide consent to participate, and 11 were excluded due to missing data.

The second cohort is a convenience sample of 578 university students recruited at the Catholic University of “Our Lady of Good Council” in Tirana (Albania), also in this case in the context of a larger cohort study, between 2016 and 2018. Six hundred and ten students were approached during lessons and invited to participate in the study. Of these, 24 did not provide consent to participate and 6 were excluded due to missing data. Questionnaires were paper and pencil and were completed at the end of lessons.

Eligible subjects provided informed consent after receiving a complete description of the study and having an opportunity to ask questions before completing the self-report questionnaires. No incentives were offered to participate. The local Ethics Committees approved all recruitment and assessment procedures.

### 2.2 Measures

All instruments used were in Italian, as the university setting in Albania is an Italian-speaking university and all of the students are fluent in Italian. The description of self-report assessment is extensively reported in another study from our group (Ciocca et al., 2017).

#### 2.2.1 Attachment styles

Attachment style was measured using the Italian version of the Relationship Questionnaire (RQ) (Bartholomew and Horowitz, 1991). The RQ is a single item measure made up of four short paragraphs, each describing a prototypical attachment style (secure, preoccupied, fearful and dismissing). Participants are first asked to choose one of the four attachment styles as the one that better describes them, providing a single item variable with four possible values. After that, participants are asked to rate their degree of correspondence to each prototype on a 7-point scale, providing four continuous variables, one for each attachment style.

For the purpose of our analysis, we used the four continuous variables only.

#### 2.2.2 Defense mechanisms

Defense mechanisms were assessed with the short form of the Italian version of the Defense Style Questionnaire (DSQ-40) (Andrews et al., 1993). DSQ is a 40 item scale on a 9-point Likert scale measuring 20 defense mechanisms grouped into three variables: Mature defenses (Sublimation, Humor, Anticipation and Suppression), Neurotic defenses (Undoing, Pseudo-Altruism, Idealization, Reaction Formation) and Immature defenses (Projection, Acting-out, Denial, Passive-Aggressiveness, Displacement, Disassociation, Splitting, Rationalization, Somatization).

#### 2.2.3 Psychological distress

Psychological distress was measured using the Italian version Symptom Check List 90-Revised Global Severity Index (SCL-90-R GSI) as the single best indicator of current psychological distress level in a non-clinical sample (Derogatis, 1992). SCL-90-R is a 90-item symptom scale that provides scores on several psychopathological domains, including anxiety, depression and psychoticism, and a global severity index that represents the average of all the 90 items.

### 2.3 Statistical Strategy

Descriptive statistics were performed on variables of interest and potential confounders. Homogeneity of the two subsamples were assessed using independent samples t-test or χ^2^test, as appropriate.

Bivariate correlations between attachment style and defense mechanisms were explored using Pearson correlation.

The influence that individual defense mechanisms and attachment style had on psychological distress, independently of each other, was assessed using linear regression. Because gender and age influenced GSI in our sample (data not shown), they were inserted as confounders in a subsequent regression model. Furthermore, because defense mechanisms undergo a maturation process from childhood to adulthood, age has been introduced as a confounding variable.

#### 2.3.1 Path analysis analytic strategy

A path model using SEM was fitted with SCL-90-R GSI as outcome variable. Based on the literature, we hypothesized an initial model in which attachment styles are exogenous variables with both direct and indirect effects through defense mechanisms on psychological distress. All variables were modelled as observed variables rather than being loaded onto a latent variable, as we were interested in discriminating the effects of each variable on psychological distress. The model was then refined inspecting individual path coefficients, Goodness-of-fit statistics, modification indices and R^2^ for SCL-90-R GSI.

An alpha error lower than 5% was set as statistical significance for all the statistical tests. All analyses were made with STATA® version 13.

## 3 Results

### 3.1 Sample

Overall sample characteristics, as well as differences in variables of interest between the two samples are reported in Table 1. In the whole sample, mean age was 22.4 (±3.02, age range 18-49) female participants outnumbered male participants (729 vs. 400; 64.6% vs. 35.4%). Mean scores of DSQ Mature, Neurotic and Immature were respectively 4.87 (1.12), 4.28 (1.23) and 3.90 (1.02). RQ Secure, preoccupied, fearful and dismissing scores were respectively 3.76 (1.80), 3.04 (1.87), 3.30 (1.99) and 3.56 (2.10).

**Table 1.** Samples characteristics and univariate comparisons between the Italian (AQ) and Albanian (TRAL) samples.

| **Variable** | **Overall**<br>**(N=1129)** | **AQ**<br>**(N=551)** | **TRAL**<br>**(N=578)** | **Statistics** | **Effect size**<br>**(*d* or *V*)** |
| --- | --- | --- | --- | --- | --- |
| *Age* | 22.4 (3.02) | 21.95 (3.4) | 22.81 (2.52) | $t = 4.83^{***}$ | $d = 0.28$ |
| *Gender* |  |  |  |  |  |
| Women | 729 (64.57%) | 390 (70.78%) | 339 (58.65%) | $\chi^2_1 = 18.14^{***}$ | $V = 0.12$ |
| Men | 400 (35.43%) | 161 (29.22%) | 239 (41.34%) |  |  |
| *Defense Mechanisms (DSQ-40)* |  |  |  |  |  |
| Mature | 4.87 (1.12) | 4.88 (1.12) | 4.64 (1.30) | $t = 3.31^{***}$ | $d = 0.19$ |
| Neurotic | 4.28 (1.23) | 4.29 (1.23) | 4.23 (1.31) | $t = 0.67$ | |
| Immature | 3.90 (1.02) | 3.90 (1.02) | 3.95 (1.10) | $t = 0.77$ | |
| *Attachment Styles (RQ)* |  |  |  |  |  |
| Secure | 3.76 (1.80) | 3.76 (1.80) | 3.93 (1.94) | $t = 1.47$ | |
| Preoccupied | 3.04 (1.87) | 3.04 (1.87) | 3.05 (1.89) | $t = 0.04$ | |
| Fearful | 3.30 (1.99) | 3.30 (1.99) | 3.35 (2.00) | $t = 0.39$ | |
| Dismissing | 3.56 (2.10) | 3.56 (2.10) | 3.95 (2.06) | $t = 3.16^{**}$ | $d = 0.18$ |
| *SCL-90-R Global Severity Index: (GSI)* | 0.71 (0.50) | 0.71 (0.50) | 0.70 (0.53) | $t = 0.11$ | |

DSQ: Defense Style Questionnaire; RQ: Relationship Questionnaire; SCL-90-R: Symptoms Check List – 90 – Revised.
\* $p < 0.05$ ; \*\* $p < 0.005$ ; \*\*\* $p < 0.001$

The two cohorts resulted homogeneous in the variables of interest except for gender proportion. An unbalance in gender proportion between the two samples was found, with more women in the Italian sample. Age mean was slightly higher in the Albanian sample (22.81 *vs*. 21.95 years). DSQ-40 Mature mean score was slightly higher in the Italian sample. RQ-dismissing mean score was higher in the Albanian sample.

### 3.2 Linear regression

Results from the regression analysis are reported in Table 2. DSQ-40 Mature, Neurotic and Immature standardized coefficients were respectively: 0.11 [0.05, 0.17]; 0.34 [0.28, 0.39]; 0.49 [0.44, 0.54]. Secure, Preoccupied, Fearful and Dismissing attachment styles standardized regression coefficients were respectively: −0.15 [−0.20, Y0.09], 0.37 [0.31, 0.42], 0.37 [0.31, 0.42] and 0.08 [0.02, 0.13]. After adjusting for age and gender, nearly no modification was observed in the regression coefficients (Table 2).

**Table 2.** Univariable Linear regression coefficients of DSQ-40 and RQ on SCL-90-R GSI.

| SCL-90-R GSI | Unadjusted Linear Regression |  | Adjusted Linear Regression <sup>a</sup> |  |
| --- | --- | --- | --- | --- |
|  | beta [95%CI] | p | beta [95%CI] | p |
| *DSQ-40* |  |  |  |  |
| Mature | 0.11 [0.05, 0.17] | <0.001 | 0.11 [0.06, 0.17] | <0.001 |
| Neurotic | 0.34 [0.28, 0.39] | <0.001 | 0.33 [0.27, 0.38] | <0.001 |
| Immature | 0.49 [0.44, 0.54] | <0.001 | 0.50 [0.45, 0.55] | <0.001 |
| *RQ* |  |  |  |  |
| Secure | -0.15 [-0.20,-0.09] | <0.001 | -0.14 [-0.20, -0.08] | <0.001 |
| Preoccupied | 0.37 [0.31, 0.42] | <0.001 | 0.36 [0.31, 0.42] | <0.001 |
| Fearful | 0.37 [0.31, 0.42] | <0.001 | 0.36 [0.30, 0.41] | <0.001 |
| Dismissing | 0.08 [0.02, 0.13] | 0.010 | 0.08 [0.02, 0.14] | 0.006 |

<sup>a</sup> Adjusted for age and gender. SCL-90-R: Symptom Check List – 90 – Revised; GSI: Global Severity Index; DSQ: Defense Style Questionnaire; RQ: Relationship Questionnaire.

### 3.3 Pairwise correlations

Pairwise correlation matrix is reported in Table 3. Pearson correlation coefficients among DSQ-40 variables were all positive, substantial and statistically significant. Correlations among RQ and DSQ variables were moderate to small (*r* between 0.05 and 0.3).

**Table 3.** Pairwise correlations among Attachment Styles and Defense Mechanisms variables.

|  | 1 | 2 | 3 | 4 | 5 | 6 |
| --- | --- | --- | --- | --- | --- | --- |
| DSQ Mature | - |  |  |  |  |  |
| DSQ Neurotic | 0.51*** | - |  |  |  |  |
| DSQ Immature | 0.49*** | 0.59*** | - |  |  |  |
| RQ Secure | 0.14*** | 0.09* | -0.01 | - |  |  |
| RQ Preoccupied | 0.13*** | 0.28*** | 0.29*** | 0.06 | - |  |
| RQ Fearful | 0.09* | 0.13*** | 0.30*** | -0.17*** | 0.35*** | - |
| RQ Dismissing | 0.16*** | 0.05 | 0.19*** | -0.08* | -0.03 | 0.24*** |

DSQ: Defense Style Questionnaire; RQ: Relationship Questionnaire. \*p<0.05; \*\*p<0.005; \*\*\*p<0.001

### 3.4 Path analysis

The initial fitted model is shown in Figure 1 and Table 4. Based on the correlational and linear regression results, the initial model was fitted excluding RQ-Dismissing and, inspecting the modification indexes, the path from RQ-Fearful and DSQ-Neurotic was excluded. All direct and indirect effects were statistically significant. In this model, the most prominent effect was from DSQ- Immature to SCL-90-R GSI, with marginal effect from DSQ-Neurotic to SCL-90-R GSI. Partial effect analysis showed that 33% and 30% of the total effect of RQ-Preoccupied and RQ-Fearful respectively were mediated. The model goodness-of-fit was sufficient: R^2^ for SCL-90-R GSI was 0.32, with an overall R^2^=0.24. In order to improve goodness-of-fit, the first model was pruned to a more parsimonious model excluding the “RQ-Secure ⇒ DSQ-Mature” path (Figure 2 and Table 5). In this model, direct and indirect effects between the retained variables and SCL-90-R and Overall R^2^ changed of a non-substantial amount. However, goodness-of-fit statistics significantly improved.

**Table 4:**
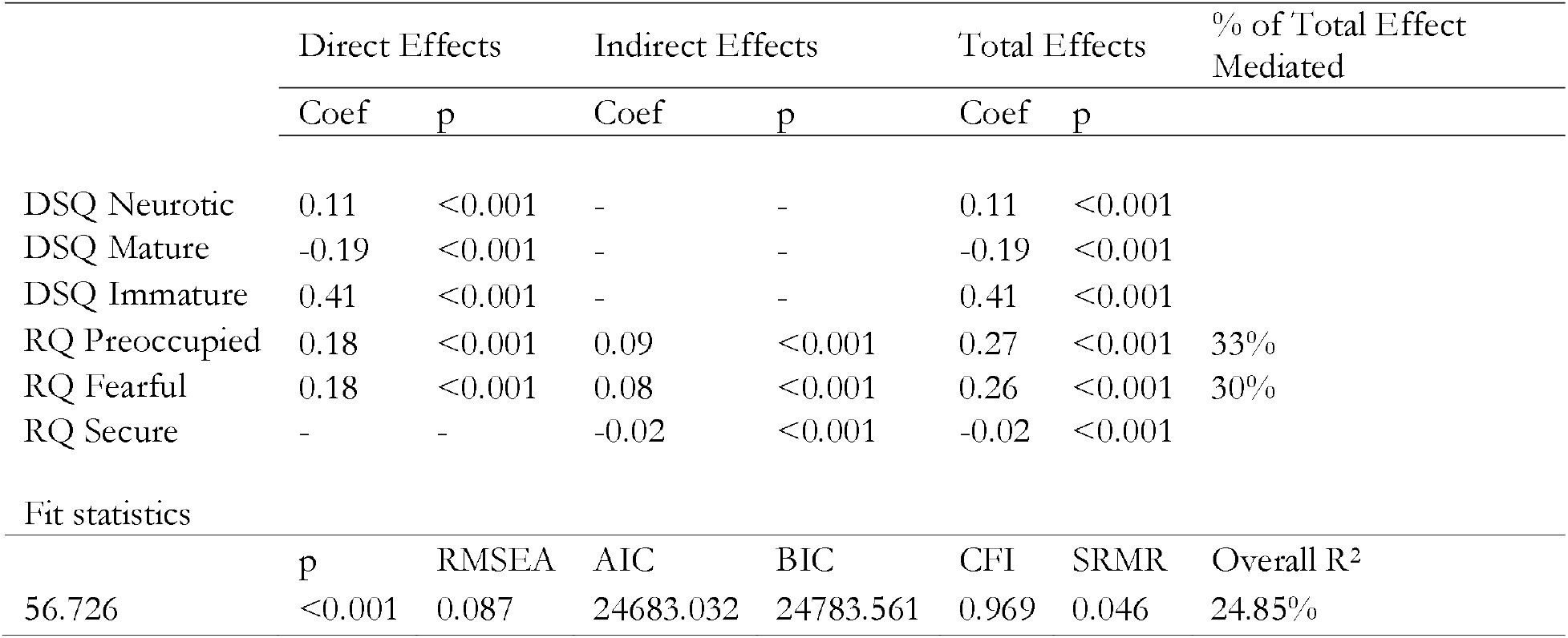
Model 1 path analysis coefficients and goodness-of-fit statistics.

**Table 5.** Model 2 path analysis coefficients and goodness-of-fit statistics.

|  | Direct effects |  | Indirect effects |  | Total Effects |  | % of Total Effect Mediated |
| --- | --- | --- | --- | --- | --- | --- | --- |
|  | Coef | p | Coef | p | Coef | p |  |
| DSQ Neurotic | 0.05 | 0.123 | - | - | 0.05 | 0.123 |  |
| DSQ Immature | 0.35 | <0.001 | - | - | 0.35 | <0.001 |  |
| RQ Preoccupied | 0.19 | <0.001 | 0.09 | <0.001 | 0.28 | <0.001 | 32% |
| RQ Fearful | 0.19 | <0.001 | 0.07 | <0.001 | 0.26 | <0.001 | 26% |
| Fit statistics |  |  |  |  |  |  |  |
|  | p | RMSEA | AIC | BIC | CFI | SRMR | Overall R<sup>2</sup> |
| 2.171 | 0.14 | 0.032 | 16940.60 | 17010.97 | 0.999 | 0.011 | 24.85% |

**Figure 1.**
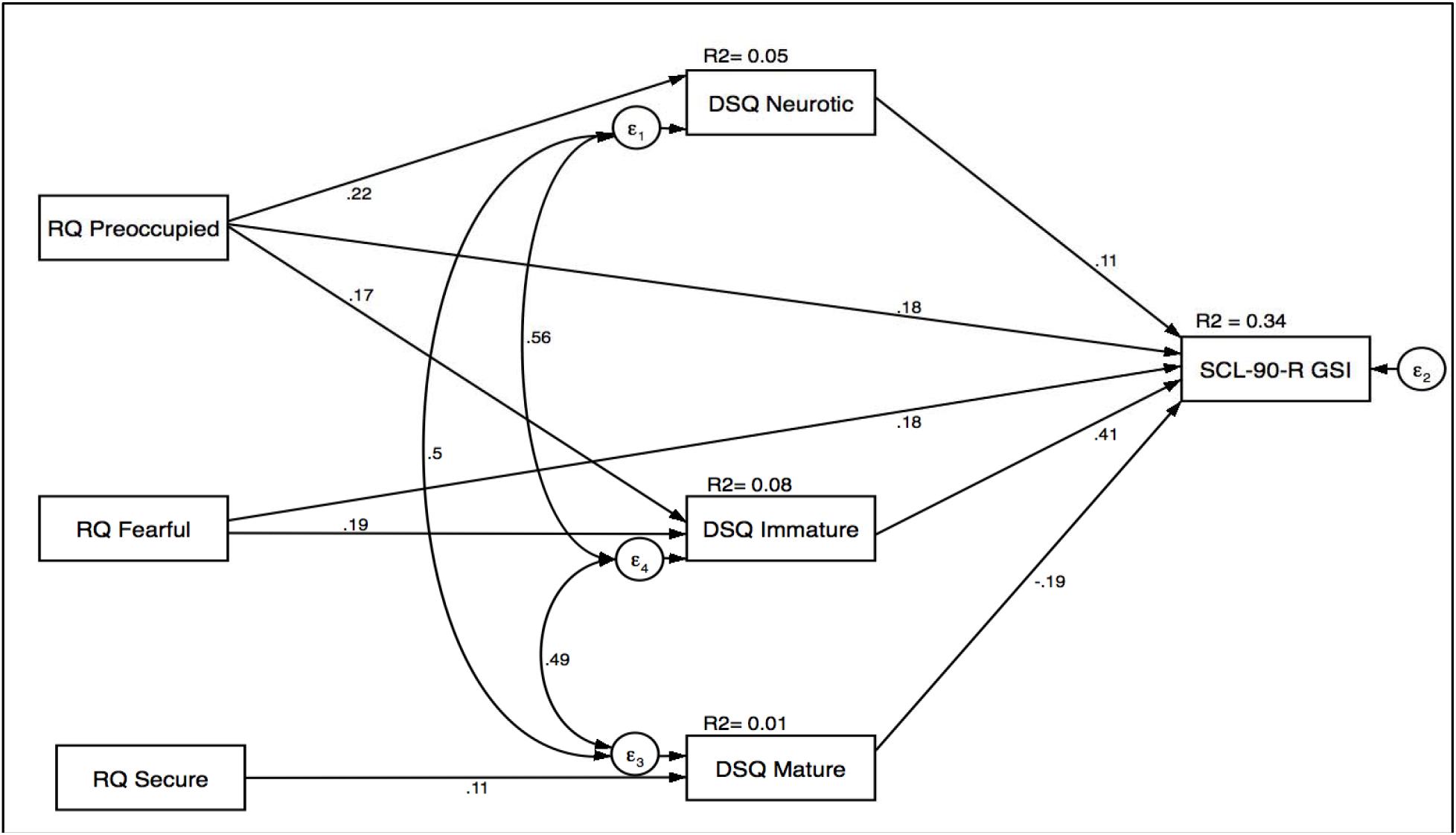
Model 1 path analysis.

**Figure 2.**
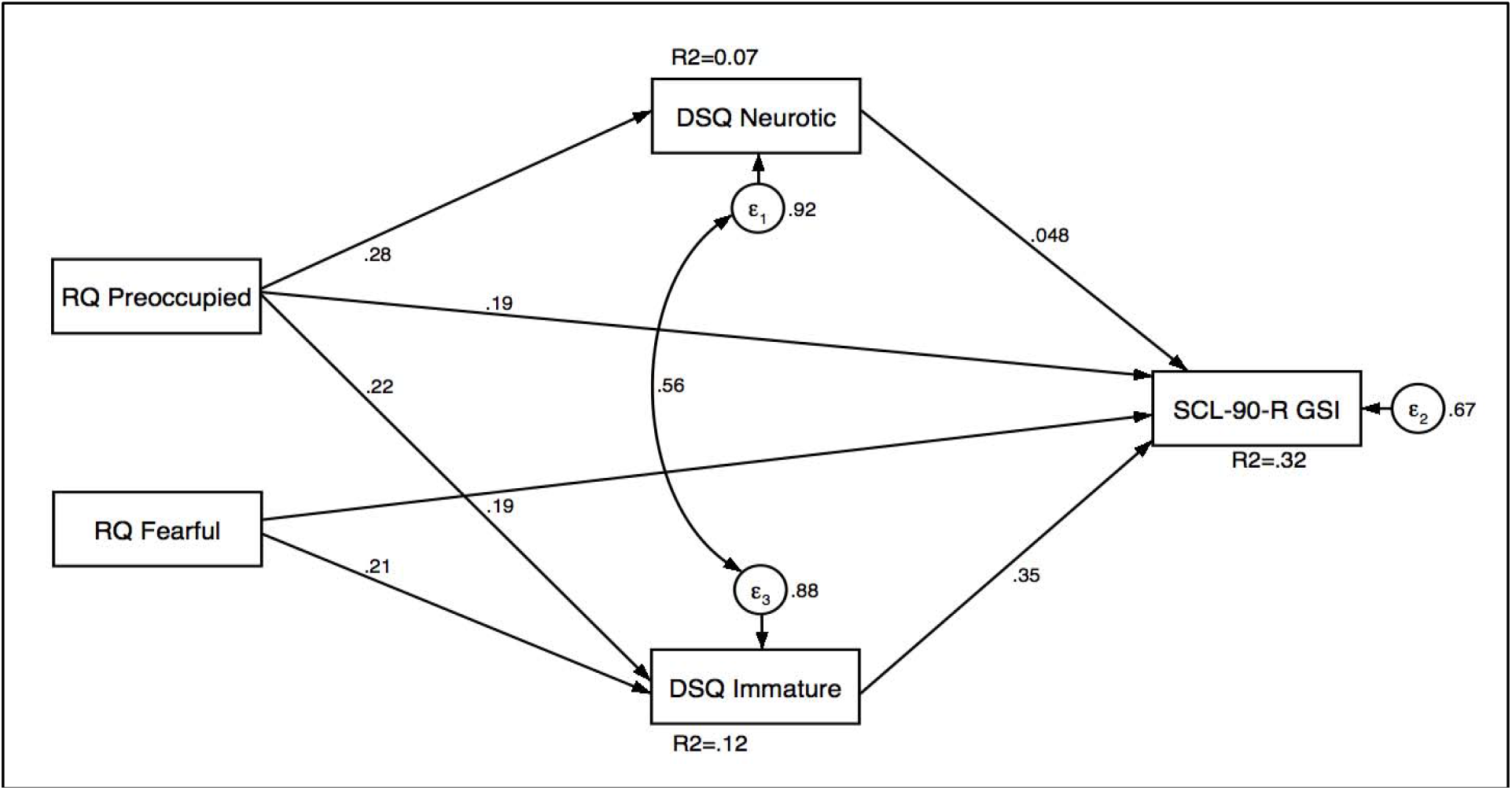
Model 2 Path analysis.

## 4 Discussion

In the present paper we investigated the influence that attachment style has on psychological distress through defense mechanisms in a large non-clinical young adult sample. To the best of our knowledge, this is the first report using path analysis to address this issue on such a large sample size, although prior works have reached considerable sample sizes as well (Laczkovics, 2018).

Our main finding is that the effect of insecure adult attachment on psychological distress is mediated by immature defenses. More in detail, we propose a model in which preoccupied and fearful attachment influence psychological distress primarily through immature defenses, while having a negligible effect through Neurotic defense mechanisms. Our model explains a substantial proportion of variance of psychological distress. Dismissing attachment showed no association with psychological distress, and was thus not included in the path model.

Additionally, we found that Secure attachment and Mature Defenses have a small but significant protective effect on psychological distress, despite worsening the overall goodness-of-fit of the model.

Our main result is coherent with previous research that associates immature defenses with insecure attachment styles (Cramer and Kelly, 2010; Mikulincer and Horesh, 1999). Considering the analytical strategy, the most similar report to ours is the work by Laczkowics (Laczkovics, 2018). In their study, the authors confirmed a mediating role of defense mechanisms between attachment style and psychological distress. The interpretation of defense mechanisms in adolescence, however, should be interpreted carefully because a certain degree of immaturity of defenses is normal during adolescence and it may not be necessarily linked to a pathological pathway. Our replication of these findings in an adult sample, as well the work by Prunas et al. on adults (Prunas, 2019) confirm and extend Laczkovics results.

Preoccupied and fearful attachment styles are both characterized by a negative self-image and high attachment anxiety, being the most relevant attachment styles related to psychological distress. However, a substantial proportion of their effect on psychological distress was mediated by defense mechanisms, suggesting that immature defense mechanisms are active in the attempt to protect a person from the distressing effect of a negative self-image. One proposed mechanism is that insecure attachment is associated with dysfunctional emotional regulation, that would in turn inhibit the development of mature defenses (Besharat, 2013). Negative self-esteem and immature defenses have been addressed as mediators of the effect of emotional abuse during childhood – which is an established harm to the attachment system, and psychological distress as adults (Finzi-Dottan and Karu, 2006). We propose that the IWM of the self is the major component of the attachment system that would drive a pathogenic effect in insecure individuals. This hypothesis would be coherent with the evidence in our results that Dismissing Attachment, which is characterized by a positive model of the self, is not associated with immature defensive mechanisms and is not associated with psychological distress. Paralleling our results with a transdiagnostic model of attachment and psychopathology proposed by Ein-Dor (Ein-Dor et al., 2016), it seems that anxious attachment styles, characterized by negative affectivity, hypervigilance to threats, and low perceived others’ responsiveness, convey the major risk for psychopathology compared to avoidant styles, characterized by cognitive and emotional avoidance and compulsive self-reliance. However, the model proposed by Ein-Dor et al., has been primarily hypothesized for clinical samples: this would mean that avoidant styles could be of importance in clinical subjects only, playing a marginal role in healthy individuals. We extend this prior evidence on a non-clinical sample, highlighting that immature defenses play an important role in determining psychological distress regardless of the presence of a clinical condition.

Regarding Secure attachment and Mature defense mechanisms, it has been suggested that the joint effect of these factors may help stabilize healthy secure individuals (Kobak and Bosmans, 2019). Our results suggest that, despite their small but still significant protective effect on general psychological distress, their explanatory gain of vulnerability to psychopathology is negligible.

### 4.1 Limitations and Strengths

Our study suffers from some limitations due to the cross-sectional design and the use of self-report psychometric instruments. Because of the cross-sectional nature of this study, the mechanistic relation between attachment style and defense mechanisms are mostly speculative. The cross-sectional design and the use of the RQ do not guarantee that attachment style as here assessed is actually antecedent to defense mechanisms in a pathway model. However, adult attachment as measured by the RQ is considered by the original authors as being loosely correlated with child attachment (Bartholomew and Horowitz, 1991), despite this point is often disputed. As usual, self-report instruments are affected by recall bias and may lead to biased results. One more limitation is the lack of assessment of the relationship status of the participants, as key constructs addressed in this study could have varied with relationship status.

Our study has some important strengths, namely the considerably large sample size and the sound analytical methodology. Path analysis suggests causal relationships between variables, that however need to be confirmed in longitudinal studies.

## 5 Conclusion

In conclusion, immature defensive mechanisms, preoccupied and fearful attachment styles are differently expressed according to the general psychological distress in non-clinical subjects. Future studies involving patients with mental disorders could help better understand the connections between insecure attachment and immature defenses with specific symptoms clusters. Therefore, these findings might induce clinicians to assess and intervene both on manifest symptoms as on defensive and relational styles, to prevent possible more severe psychological distress conditions in non-clinical subjects, particularly.

## Data Availability

data available upon request from the authors

## Acknowledgements

The authors would like to thank all the University students who participated in this study.

## Declarations of interest

None.

## Role of the funding source

This research did not receive any specific grant from funding agencies in the public, commercial, or not-for-profit sectors.

## Notes

### Competing Interest Statement

The authors have declared no competing interest.

### Funding Statement

no funding was provided for this study

